# Free-Living Hip Accelerometry Predicts Frailty Status and 12-Month Frailty Decline in Older Adults

**DOI:** 10.1101/2025.07.09.25330372

**Authors:** Benjamin Kramer, Yanan Long, Michelangelo Pagan, Sylvia Brown, Andrey Rzhetsky, Megan Huisingh-Scheetz

**Author notes:** Co-first authors (Benjamin Kramer, Yanan Long).

## Abstract

Frailty is a clinical syndrome in older adults characterized by heightened vulnerability to adverse outcomes, yet it remains under-assessed in routine practice due to time-intensive evaluation methods. Wearable accelerometers offer a promising approach for passive, continuous frailty monitoring. We analyzed data from 151 community-dwelling older adults (≥65 years) in the FACE Aging Study who wore hip accelerometers for seven days. Machine learning models were developed to classify baseline frailty status using an adapted frailty phenotype and predict 12-month frailty decline. Features derived from accelerometry data included sleep patterns, activity levels, sedentary behavior, and circadian rhythms. Multiple algorithms were compared, including deep neural networks (ResNet), gradient boosting machines (LightGBM, XGBoost, CatBoost), and multilayer perceptrons (MLPs). For baseline frailty classification, LightGBM achieved the highest performance with an F1 score of 0.898, precision of 0.815, perfect recall (1.000), and AUROC of 0.887. For predicting 12-month frailty decline, multilayer perceptron performed best with an F1 score of 0.812, precision of 0.891, and recall of 0.746. Key predictive features included low overall activity, high sedentary behavior, fragmented activity patterns, and poor sleep quality, which aligned with established frailty phenotypes. One week of free-living hip accelerometry data can accurately identify current frailty status and predict short-term frailty progression in older adults. These findings support the feasibility of developing automated, scalable frailty screening tools that could enable proactive interventions and transform geriatric care from reactive to preventive approaches. External validation in larger, diverse populations is needed to confirm generalizability.

## Introduction

Frailty in older adults is a clinical syndrome marked by heightened vulnerability to adverse outcomes due to diminished physiologic reserve, particularly when faced with stressors [6]. It is diagnosed by a constellation of features: exhaustion, low physical activity, low strength, unintentional weight loss, and slow gait speed [6]. These measures include a mix of self-reported and physical performance assessments. Presence of 1-2 of these features defines pre-frailty, an earlier stage of the syndrome, while greater than 3 of these features indicate frailty [6]. These clinical signs reflect an underlying deterioration in physiologic processes, leaving an individual less able to cope with stressors [8]. Consequently, frail older adults are at a higher risk of disability, hospitalization, and mortality when faced with major stressors such as acute illness, surgery, or chemotherapy [6]. Evidence suggests that targeted interventions including resistance exercise training, protein supplementation and specialization, comprehensive geriatrics care can improve frailty outcomes. However, frailty is under-assessed in routine practice, in part because standard assessments (e.g. Fried’s phenotype or Clinical Frailty Scale) require time and special training and are rarely performed during a short clinic visit [6,8]. Thus, there is a need for more the implementation of scalable, passive frailty screening tools to identify patients at risk for frailty and to trigger preventative interventions.

Wearable sensors such as accelerometers offer a promising approach to monitor frailty [3,9,20,21]. Accelerometers are lightweight, noninvasive devices which passively and continuously record movement at high frequency in free-living conditions [2,14]. From the raw acceleration signal, one can derive a rich set of measures of daily activity, sedentary behavior, sleep patterns, and circadian rhythms [2,5,14]. These patterns contain critical information about an individual’s functional status and the progression of aging-related decline [18,19]. Accelerometers can produce a sub-second resolution of 24-hour activity-inactivity-sleep movement patterns [2,5]. These various acceleration patterns reveal critical information about aging pathologies and individual-specific pace of frailty onset.

The major unaddressed clinical needs in the care for older adults include (1) identifying patients who are most at risk of becoming pre-frail or frail; and (2) to flag patients who are at the highest risk of an accelerated, short-term frailty decline. In this study, we leverage continuous hip-worn accelerometry data from a cohort of community-dwelling older adults to (1) classify baseline frailty status and (2) prospectively predict 12-month frailty decline using machine learning. We focus on a single observational cohort with hip accelerometer monitoring, the Frailty, Aging, Body Composition and Energy Expenditure in Aging (FACE) study. By using free-living accelerometer measurements as inputs to advanced models – including deep neural networks and gradient boosting machines – we aim to identify individuals at risk of transitioning from robust to frail states within one year. Our approach requires no active patient input beyond wearing a device, making it a practical tool for widespread frailty screening. We hypothesize that patterns in daily movement and sleep, captured by hip accelerometry, can serve as early warning signs of frailty progression. We present results demonstrating that wearable-derived metrics can accurately distinguish frailty phenotype categories at baseline and forecast which older adults will experience worsening frailty over a 12-month period.

## Methods

### Study Cohorts and Design

The FACE Aging Study recruited community-dwelling, older adults ⩾ 65 years (n = 151) from the community around the University of Chicago. Exclusion criteria included hospitalization, surgery or procedure within two months of participating in the study; new or change in dose of thyroid (levothyroxine) or a diuretic (furosemide, hydrochlorothiazide, or spironolactone) medication within two months of participating in the study; use of oral steroids; use of beta-blocker (e.g., metoprolol, atenolol, or carvedilol); persistent hyperglycemia greater than 250; life expectancy less than one year; and history of moderate or advanced dementia or a Montreal Cognitive Assessment (MoCA) score ⩾ 18.

Hospital, surgery, medication, and hyperglycemia exclusion criteria were required to optimize resting metabolic rate testing at baseline, data that were not used in the current analyses. Data collection occurred at baseline with a survey and physical exam in the clinic, followed by a seven-day free-living hip accelerometry wear period. Fasting resting metabolic rate measurement with indirect calorimetry and DEXA scan for body composition were then conducted within 2 weeks of the baseline assessment. At one-year, data collected occurred with a follow-up survey and physical exam in the clinic. We restricted the study sample to participants with complete covariate data and one or more valid (⩾ 10 daytime hours) accelerometer-wear days.

### Accelerometer data processing

Following the baseline survey and physical exam, all participants were asked to wear an Actigraph wGT3X+ hip accelerometer over the mid, anterior right hip. The device was placed in the clinic and secured with an elastic belt. Study participants were asked to wear the device continuously for 7 full days (including during bathing or showering) but actual compliance may vary. Data were collected at a frequency of 30 Hz.

The hip accelerometer data were extracted from the devices as gt3x files using the ActiLife software (version #, year, ActiGraph, Inc). The gt3x files contained the raw acceleration data (gravitational units) in each of the three axes for each timestamp (i.e., 30Hz = 30 timestamps per second). The gt3x files were then opened by the Python-language package pygt3x (Neishabouri et al., 2022) created by ActiGraph.

The files were calibrated to account for gravity using the same package pygt3x (Neishabouri et al., 2022). There were four individuals who had 1 second of the entire wear period that had duplicate accelerations recorded for each timestamp (i.e. 30 Hz timestamps with more than 1 reading). Only unique recordings were retained in the file for a given timestamp. The files were then converted to tab-separated text files. Missing data in each axis (x, y and z) were imputed using the median of the non-missing data for the remainder of the entire time series (i.e., 7-day wear period). In this sample, ## percent of the data were missing and imputed. These imputed time series tab-separated text files were used to generate the below measures.

Subsequent analyses proceeded in two processing pathways. 1) First, the metrics Euclidean norm minus one (ENMO) and the related ENMO/minute; the vector magnitude count (VMC) and related VMC/minute; and the Actigraph “counts” were calculated using the imputed time series tab-separated text files. ENMO/minute, VMC/minute and Actigraph counts were used to generate the activity, sleep and harmonic variables.

The ENMO was calculated as

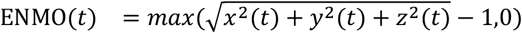

where *t* is in the unit of 1/30 seconds. Then, the ENMO time series data were aggregated into non-overlapping sliding windows (“epochs’’) with a length of 60 seconds across the entire wear period. The ENMO values within each epoch were then averaged as a new time series 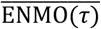

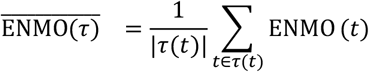

where *τ* indexes the epoch within each day of wear and *τ*(*t*) is the epoch corresponding to the time t. The VMC times series were calculated as the absolute value of the difference between ENMO(*t*) and 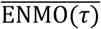 time series:

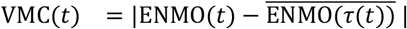

From this data, the 60s epoch-level average 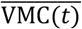 was calculated in the similar way as 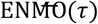:

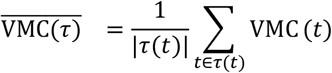

Finally, a set of statistical and harmonic features was calculated for each of 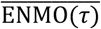 and 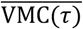 using the 60s epoch-level averages of ENMO and VMC as in previous work (Table 5 of Shi et al. (2022)): 98 accelerometry harmonic features (e.g. entropy, skewness, kurtosis, fast Fourier transformations, amplitude).

Next, the Sadeh algorithm [17] was applied to the Actigraph counts time series calculated as [14], each epoch was identified as sleep or awake. Then, using the modified Tudor-Locke method [22] (12 minutes instead of 10 minutes in detecting wake time), in-bed intervals were identified, which enabled the calculation of (1) total sleep time calculated as the sum of all epochs spent in sleep; (2) wake time identified as the average time of end of sleep and start of daytime interval (3) wake after sleep onset calculated as the sum of the epochs classified as awake during the sleep interval; (4) sleep fragmentation calculated as the percentage of epochs with motion during the sleep interval; (5) percentage sleep calculated by dividing sleep minutes by total sleep interval minutes. Moreover, mobility measures were calculated during the awake interval: (1) sum, average and 99th percentile of ENMO during the awake interval; (2) time spent in various intensities of activity levels define as the average daily time spent in minutes of light, moderate, or vigorous activity using established count thresholds [12]; (3) average number of sustained moderate-to-vigorous active bouts calculated as the mean daily number of bouts lasting ⩾ 5 minutes in duration above moderate intensity count thresholds; (4) transition probabilities from active to sedentary periods (and vice versa) defined as the reciprocal of average bout lengths per day [18]. The resulting features of this branch were collectively designated Tabular data. See Table 1 for a summary of the features.

**Table 1:**
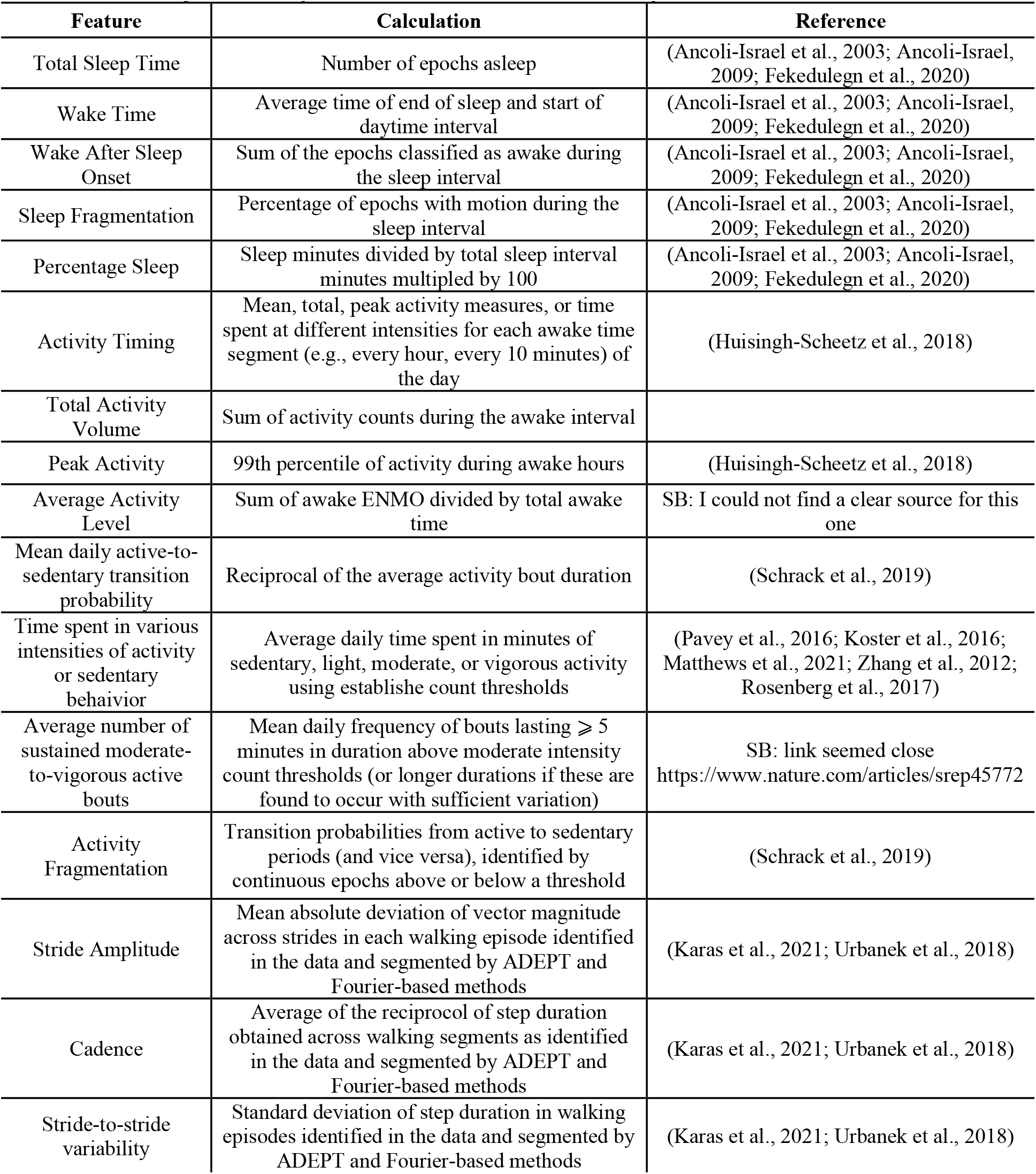
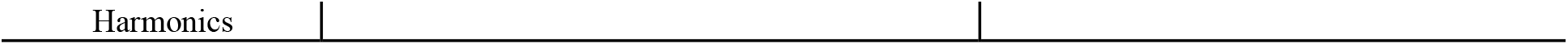
Sleep and Activity Features Derived from Accelerometry Data.

**Table 2:** Summary statistics for demographic variables, q_p_ is the p^th^ percentile.

|  | **robust** | **pre-frail** | **frail** | **total** |
| --- | --- | --- | --- | --- |
| **Age** |  |  |  |  |
| **Mean** | 71.24 | 74.37 | 72 | 72.72 |
| **Standard deviation** | 5.74 | 6.32 | 4.41 | 6.049 |
| **$Q_0$** | 65 | 65 | 65 | 65 |
| **$Q_{50}$** | 69 | 73 | 72 | 71 |
| **$Q_{100}$** | 90 | 89 | 77 | 90 |
| **Education** |  |  |  |  |
| **Less than High School** | 1 | 1 | 0 | 2 |
| **Greater than or Equal to High School** | 41 | 41 | 8 | 90 |
| **Race** |  |  |  |  |
| **Black or African American** | 29 | 35 | 8 | 72 |
| **White or Other** | 13 | 6 | 0 | 19 |
| **Monthly Income** |  |  |  |  |
| **0 – 1,999** | 19 | 18 | 4 | 41 |
| **2,000 – 3,999** | 10 | 14 | 4 | 28 |
| **4,000 – 5,999** | 5 | 5 | 0 | 10 |
| **>6,000** | 4 | 4 | 0 | 8 |
| **Mean** | 3,040 | 2,734 | 1,565 | 2,760 |
| **Standard deviation** | 3,503 | 2,198 | 1,186 | 2,794 |
| **$Q_0$** | 0 | 0 | 0 | 0 |
| **$Q_{50}$** | 1,900 | 2,300 | 1,386 | 2,000 |
| **$Q_{100}$** | 19,000 | 12,000 | 3,500 | 19,000 |

### Measures

Frailty was assessed using an adapted frailty phenotype in both cohorts. In the hip accelerometry cohort, five measures were used to construct the adapted frailty phenotype. Unintentional weight loss was identified by calculating the difference between measured baseline weight and self-reported weight 1 year prior; those with greater than five percent of body weight or ten pounds lost were assigned a point. Weakness was assessed using the average of three dominant grip strength measurements (Jamar hydraulic hand dynamometer); those who had strength performance below the established body mass index- and gender-adjusted cut-points were assigned a point [6]. Slowness was assessed using the average of three, 15-foot usual walk times; those with usual walk times below gender- and height-adjusted cut points were assigned a point [6]. Exhaustion was assessed using two self-reported survey questions. Participants reported the frequency they “felt that everything was an effort’’ or that they “could not get going’’ in the prior week 3-4 days (response options: None of the time, Some or little of the time (1-2 days), A moderate amount of the time (3-4 days), or Most of the time (>4 days)). Those reporting having either feeling a moderate amount of the time were assigned a point. Finally, low activity was assessed using the kcal/week calculated from responses to the 6-item Minnesota Leisure-Time Physical Activity Questionnaire. Those reporting physical activity levels below gender-adjusted thresholds were assigned a point [4]. Points were summed to a total score ranging from zero to five: zero points indicated a non-frail status, one to two points indicated a pre-frail status, and three or more points indicated a frail status.

Physical frailty was measured at baseline and 1 year.

## Results

### Baseline Frailty Classification Performance

Machine learning models demonstrated strong performance in classifying baseline frailty status using one week of hip accelerometry data. Table 3 presents the performance metrics for distinguishing between frail/pre-frail versus robust participants at baseline. The best-performing model was LightGBM, achieving an F1 score of 0.898, precision of 0.815, perfect recall (1.000), accuracy of 0.887, and AUROC of 0.887. ResNet also performed well with an F1 score of 0.840 and balanced precision (0.842) and recall (0.838). XGBoost achieved comparable performance (F1 = 0.804) with perfect recall but lower precision (0.673). Traditional tree-based methods showed more modest performance, with DART achieving an F1 score of 0.761. The multilayer perceptron (MLP) showed high precision (0.933) but poor recall (0.410), resulting in a lower overall F1 score of 0.570. CatBoost achieved moderate performance across all metrics (F1 = 0.624).

**Table 3:** Best-performing model in terms ofF1score for binarized baseline (0-month) frailty score.

| Method | $F_1$ (↑) | Precision (↑) | Recall (↑) | Accuracy (↑) | AUROC (↑) |
| --- | --- | --- | --- | --- | --- |
| *Tabular* |  |  |  |  |  |
| CatBoost | 0.624 | 0.690 | 0.570 | 0.657 | 0.657 |
| LightGBM | 0.898 | 0.815 | 1.000 | 0.887 | 0.887 |
| XGBoost | 0.804 | 0.673 | 1.000 | 0.757 | 0.757 |
| DART | 0.761 | 0.697 | 0.837 | 0.736 | 0.736 |
| MLP | 0.570 | 0.933 | 0.410 | 0.690 | 0.690 |
| ResNet | 0.840 | 0.842 | 0.838 | 0.840 | 0.840 |

### Longitudinal Frailty Decline Prediction

Table 4 shows the performance of machine learning models in predicting 12-month frailty decline (defined as an increase in frailty phenotype score from baseline to follow-up). For prospective frailty decline prediction, ensemble boosting methods demonstrated superior performance. MLP achieved the highest F1 score of 0.812 with good precision (0.891) and recall (0.746). CatBoost performed comparably with an F1 score of 0.807, precision of 0.964, and recall of 0.694. XGBoost also showed strong performance (F1 = 0.796) with high precision (0.856) and moderate recall (0.743). DART achieved notable performance with the highest recall (0.902) but lower precision (0.645), resulting in an F1 score of 0.752. LightGBM, despite its excellent baseline classification performance, showed more modest longitudinal prediction ability (F1 = 0.704). ResNet, which performed well for baseline classification, showed the poorest performance for longitudinal prediction (F1 = 0.589) with low recall (0.448).

**Table 4:** Best-performing model in terms ofF1 score for binarized difference between 12 months in frailty score.

| Method | $F_1$ (↑) | Precision (↑) | Recall (↑) | Accuracy (↑) | AUROC (↑) |
| --- | --- | --- | --- | --- | --- |
| *Tabular* |  |  |  |  |  |
| CatBoost | 0.807 | 0.964 | 0.694 | 0.834 | 0.834 |
| LightGBM | 0.704 | 0.891 | 0.582 | 0.756 | 0.756 |
| XGBoost | 0.796 | 0.856 | 0.743 | 0.809 | 0.809 |
| DART | 0.752 | 0.645 | 0.902 | 0.703 | 0.703 |
| MLP | 0.812 | 0.891 | 0.746 | 0.828 | 0.828 |
| ResNet | 0.589 | 0.857 | 0.448 | 0.687 | 0.687 |

### Model Performance Comparison

The results demonstrate distinct patterns in model performance between cross-sectional classification and longitudinal prediction tasks. For baseline frailty classification, deep learning approaches (ResNet) and gradient boosting methods (LightGBM, XGBoost) achieved the strongest performance, with F1 scores ranging from 0.804 to 0.898. For longitudinal frailty decline prediction, ensemble methods (MLP, CatBoost, XGBoost) consistently outperformed other approaches, with F1 scores ranging from 0.796 to 0.812. Notably, the models achieving the highest recall for decline prediction (DART with 0.902 recall) may be particularly valuable for clinical screening applications where sensitivity to detect at-risk individuals is prioritized. All top-performing models achieved AUROC values above 0.75 for both tasks, with the best baseline classification models reaching AUROC values approaching 0.90, indicating strong discriminative ability for identifying frailty status and predicting decline using accelerometry-derived features.

## Discussion

In this study, we demonstrated that wearable sensor data from a hip accelerometer can be used to both assess current frailty and forecast future frailty progression in community-dwelling older adults. Using one week of free-living accelerometry from the FACE cohort, our machine learning models classified baseline frailty status with high accuracy and predicted 12-month frailty worsening. To our knowledge, this is the first study to successfully predict short-term frailty decline using continuous free-living activity monitoring [3,9,18,19]. These findings have significant implications for developing non-invasive, automated frailty screening tools.

We found that certain activity patterns including low overall activity level, high sedentary behavior, fragmented activity bouts, and poor sleep quality were strongly associated with frailty and its progression [1,2,6,18]. These patterns are intuitively consistent with the frailty phenotype, which includes fatigue and reduced activity [6]. Our models effectively captured these signals. Notably, advanced machine learning methods improved prediction performance over simpler approaches. In cross-section, a deep residual network slightly outperformed other models, suggesting that complex nonlinear feature interactions (and possibly higher-order feature combinations) contributed to identifying frailty.

In the longitudinal prediction, ensemble methods (LightGBM, CatBoost) achieved the best balance of recall and precision. The difference in “best” model between tasks may reflect the nature of the prediction problem: distinguishing frail vs. robust at a single time point may benefit from deep pattern recognition (for example, capturing subtle daily activity rhythms), whereas predicting change over time may rely on a combination of risk factors that boosting algorithms can weight effectively. Nonetheless, performance differences between ResNet and LightGBM were small, and all top models (including the MLP) performed in a narrow band. This convergence increases confidence that the results are not an artifact of one particular algorithm but rather driven by true underlying relationships between accelerometry features and frailty outcomes.

The ability to forecast frailty progression has significant clinical value. Frailty is best managed when detected early with interventions such as tailored exercise programs, nutritional supplementation, and preventive geriatric care can slow or reverse frailty in its initial stages [6,8]. If our algorithm flags a robust older adult as high-risk for becoming pre-frail in the next year, clinicians could proactively engage them in interventions (e.g. resistance training or physical therapy) before overt frailty manifests. Similarly, patients identified as currently pre-frail could be monitored more closely or referred to geriatric specialists to prevent further decline. Another application is in pre-surgical evaluation: accelerometer-based frailty assessment could help identify older surgical candidates who are frail or at risk of decompensation, allowing for prehabilitation or altered management to reduce post-operative complications [7]. Overall, this technology could enable a shift from reactive to proactive care in aging and detecting risk early and intervening to maintain function.

This study has several limitations. First, our sample size was modest (N~90 for baseline analysis), drawn from a single-center cohort. While we achieved strong cross-validated performance, external validation in a larger, independent population is needed to ensure generalizability. We focused on one specific cohort (FACE study) with hip-worn devices; thus, results may not directly translate to other wearables (e.g., wrist devices) or different populations [3,11,16]. Indeed, prior research indicates some differences in hip vs. wrist accelerometers and their relation to physical performance [3,11]. Second, our follow-up period was one year. While clinically relevant, it is relatively short; some individuals categorized as non-decliners in that interval might decline over a longer period. We did not have 5-year accelerometry follow-up data in this cohort to evaluate longer-term predictions. Relatedly, we defined frailty decline as an increase in the frailty phenotype score. It is possible some participants’ frailty status worsened in ways not captured by the phenotype (e.g., new comorbidities) or even fluctuated and improved, but our binary outcome did not differentiate those nuances. Third, although we included many features, there may be other sensor-derived metrics (e.g. heart rate variability from wearables, or contextual information about where activity occurs) that could further improve predictions [10,13,20]. We also did not directly incorporate physiological or blood-based biomarkers, which have been linked to frailty in other studies – our goal was to see how far we could get with wearables alone. In practice, a multi-modal model combining accelerometry with key clinical data might yield even better accuracy, though at the expense of simplicity. Finally, we acknowledge that our modeling approach, while sophisticated, is not fully transparent. Gradient boosting and neural nets operate as “black boxes” to an extent. We did derive feature importance rankings from the boosting models, which suggested that high-level features like total activity, moderate activity time, and sleep fragmentation were top predictors (lending face validity to the models). Future work could employ explainable AI techniques (e.g. SHAP values) to further interpret how specific activity patterns influence the frailty risk prediction for an individual. This could enhance clinician trust and provide personalized insights (for example, informing a patient that “low afternoon activity and very irregular sleep times are contributing to your frailty risk”).

## Conclusion

In conclusion, our study shows that one week of free-living hip accelerometer data can accurately identify older adults’ frailty status and even anticipate short-term frailty progression. Wearable sensors capture objective signatures of diminished physiologic reserve including low physical activity, fragmented activity rhythms, and poor sleep which correspond closely to the frailty construct [1,2,6,18]. Leveraging these signals, machine learning models (especially ResNet deep networks and LightGBM/CatBoost boosters) achieved high predictive performance, matching or exceeding prior more invasive approaches. These findings support the feasibility of a “digital frailty index” that could be implemented at scale for early detection. With further validation, such models could be deployed in smartphones or wearable devices to continuously monitor at-risk seniors and alert healthcare providers to early signs of frailty.

Proactive interventions could then be initiated, potentially improving outcomes by maintaining independence and preventing downstream adverse events. Our work contributes to the growing evidence that passive sensing technology can transform care for older adults by enabling timely, personalized geriatric risk assessment [3,9,13,19]. Future studies will expand on this foundation by testing our accelerometry-based frailty predictions in larger and more diverse cohorts, exploring longer-term forecasting (e.g. 3–5 year frailty outcomes), and integrating additional data streams to further enhance accuracy.

## Data Availability

All data produced in the present study are available upon reasonable request to the authors

## Ethics Statement

The study was approved by the University of Chicago Institutional Review Board (IRB #13-0443 and IRB #11-0205 for the hip accelerometry study). Study participants provided written informed consent.

## Notes

### Competing Interest Statement

The authors have declared no competing interest.

### Funding Statement

NIH/NIMH award 1R01MH137646-01

